# Prevalence and determinants of not testing for HIV among young adult women in Papua New Guinea: findings from the 2016–2018 Demographic and Health Survey

**DOI:** 10.1101/2023.05.07.23289638

**Authors:** McKenzie K. Maviso, Fatch Welcome Kalembo

**Affiliations:** School of Medicine and Health Sciences, University of Papua New Guinea, Port Moresby, Papua New Guinea; School of Nursing, Curtin University, Perth, Western Australia, Australia

## Abstract

**Objective:** The study investigated the factors associated with not ever testing for HIV among women aged 15–29 years in Papua New Guinea (PNG).

**Design and setting:** The study used secondary data from the 2016–2018 Demographic and Health Survey (DHS) of PNG, a nationally representative cross-sectional survey that used a two-stage stratified sampling.

**Participants:** A total weighed sample of 5,164 young adult women aged 15–29 years were included in the study.

**Primary outcome measure:** Ever been tested for HIV was the primary outcome of the study. All analyses were adjusted using survey weights to account for unequal sampling probabilities.

**Results:** The prevalence of not testing for HIV was 58.8% (95% CI: 57.4%, 60.1%). The mean age was 21.65 (SD ± 4.23) years. The majority (62.9%) of young adult women in rural areas were not tested for HIV. In the multivariable analysis, those who were never married (AOR: 4.9, 95% CI: 3.6–6.6), had poor wealth index (AOR: 1.8, 95% CI: 1.3–2.5), from rural areas (AOR: 2.0, 95% CI: 1.5–2.6), from the Momase region (AOR: 1.3, 95% CI: 1.0–1.7, did not read newspaper or magazine (AOR: 1.7, 95% CI: 1.3–2.1), did not listen to radio (AOR: 1.5, 95% CI: 1.1–2.0), experienced early sexual debut (AOR: 1.5, 95% CI: 1.1–1.9), had one sexual partner (AOR: 1.5, 95% CI: 1.2–2.0), and reported no STI in the past 12 months (AOR: 1.8, 95% CI: 1.1–3.1) had higher odds of not testing for HIV.

**Conclusions:** Our study found a very high unmet need for HIV testing among young adult women in PNG. Health promotion programmes should be designed to increase HIV knowledge and access to HIV testing services. Furthermore, efforts to optimise HIV testing services should target women who are disadvantaged and from rural areas.

**Strengths and limitations of this study:**

- This study used a nationally representative large sample of young adult women aged 15–29 years from the 2016–2018 PNGDHS; thus, the findings are generalisable to the entire population of this age group.
- The study provides much-needed data for strategic planning and programming to improve the health outcomes of young adult women in the country.
- This is a cross-sectional study, and as such, it is not possible to make causal inferences.
- The level of knowledge or awareness of where to get tested for HIV was not included in the analysis, and it remains unknown whether this variable contributes to the high proportion of respondents who remain unaware of their HIV status.
- Responses to the questions from respondents were self-reported, and this could have resulted in behavioural desirability bias.

## Introduction

Globally, an estimated 39 million people were living with HIV in 2022, and approximately 17% (6.5 million) of these were from the Asia-Pacific region. [1] Among people living with HIV, young women continue to endure a disproportionate burden of the virus. [2] Young women are twice as likely as young men of the same age to have HIV; [3] are frequently unaware of their HIV status and lack access to antiretroviral therapy (ART) and care. [4] They are more likely to experience physical or sexual violence from intimate partners compared to their male counterparts. [5,6] The lifetime prevalence of intimate partner violence among young women is 16%. [6,7] The violence experienced by young women is a significant factor in causing unequal infection rates in settings with high HIV prevalence. [8] Such disproportionately high rates of HIV infection in young women compared to young men are illuminated by an array of influences. For instance, social and institutional norms (e.g., male superiority) and gender-based power inequalities (e.g., sexual entitlement within relationships) prevent women from negotiating safe sex practices, which frequently impact their access to sexual and reproductive health care and HIV prevention services. [9–12]

Voluntary counselling and testing (VCT) for HIV is a gateway to HIV treatment and care for those who test positive and a prevention strategy for those who test negative. [13] Awareness of one’s HIV serostatus is associated with risk reduction and behaviour change. [14,15] Newly diagnosed HIV individuals can be promptly connected to ART, care, and support services. Timely HIV diagnosis and treatment access are crucial for preventing transmission, as newly diagnosed individuals can immediately be linked to ART, care, and support services. [16]

While access to HIV testing, treatment, and support services has been expanding globally, the response to HIV testing has remained inadequate, particularly among young women who are recognised for having poor health-seeking behaviours in highly prevalent settings. [17,18] Factors associated with low coverage or not testing for HIV are multifactorial and include the fear of HIV test results and stigma, the perceived low risk of contracting HIV, and the psychological burden of living with HIV. [17,19] In addition, knowledge and attitude towards HIV testing services, age, education, engagement in sexual relationships, and distance to access testing facilities are associated with low uptake of HIV testing among young people. [20,21]

PNG is one of the most HIV-affected countries in the Western Pacific Region, with a high HIV prevalence projected at 0.9% in the general adult population. [22,23] In 2022, an estimated 72,000 people were living with HIV in PNG. [24] In addition, women of childbearing age (15–49 years) in PNG represent a disproportionate population that requires adequate HIV prevention services, including sexual and reproductive health care. [25–27] In 2022, women accounted for about 67% (41,000) of HIV, with 3,600 new HIV infections in the country. [24] It has been reported that HIV prevalence among PNG women is attributed to multiple factors such as gender inequality, sociocultural norms, household poverty, and unsupported social and structural factors, including unequal access to sexual and reproductive health services, increasing women’s vulnerability to HIV infection. [28–31]

To address the burden of HIV, the government of PNG has integrated the WHO-recommended VCT for HIV into the country’s critical HIV prevention and control strategies. [32] With more ART programmes available, efforts to strengthen HIV testing have become increasingly important for early diagnosis and linking access to treatment, adherence to treatment, and viral load suppression. [32,33] Despite the country’s efforts to reduce HIV transmission, VCT uptake, linkage to ART, and support services remain problematic. [33–35] In particular, young women’s awareness of HIV prevention strategies, such as consistent condom use and reducing the number of sexual partners, remains low (24%). [2] For this study, young adult women aged 15–29 years were included in the analysis. Previous studies have investigated sexual, social, and structural determinants of risk behaviours and adverse health outcomes, including HIV acquisition, among this group (aged 15–29). [36–38] However, research on factors associated with not testing is limited, especially in young women, who face a high risk of HIV. Furthermore, there have been no studies investigating how these factors influence HIV testing, an integral component of an HIV prevention package. Therefore, this study aimed to examine factors associated with not testing for HIV among young adult women in PNG. Recognising and addressing the challenges that young women face in accessing HIV testing services is critical to achieving the ambitious UNAIDS 95-95-95 targets to end the HIV/AIDS epidemic. [39]

## Methods

### Study design and sampling

This study is a result of the analysis of the 2016–2018 PNGDHS data, which is a nationally representative cross-sectional survey that provides data to monitor and evaluate population, health, and nutrition programmes. The PNGDHS researchers used a two-stage stratified sampling technique to recruit participants for the study. The first stage involved selecting 800 census units (CUs), which was achieved through a probability proportional to CU size. The second stage involved the systematic selection of 24 households from each cluster (CU) through equal probability sampling, with the resulting sample consisting of 19,200 households. The survey used the CUs from the 2011 PNG National Population and Housing Census, which used structured sampling. Additionally, PNG has 22 administrative provinces, and each province is subdivided into urban and rural areas. Each province also has districts, and each district is divided into local-level government wards, where each ward is comprised of several CUs. The PNG National Statistical Office (NSO) and ICF International collected the data. In the interviewed households, 18,175 women aged 15–49 years were identified for individual interviews. Of these, 15,198 women participated in the survey successfully, yielding a response rate of 84%. Among those who were interviewed successfully, 5,164 (the weighted sub-sample) were young adult women aged 15–29 years who had already initiated sex and were included in this study. Information about the methodology, pretesting, and training of field officers, the sampling design, and sample selection is available in the PNGDHS final report [40] and online at https://dhsprogram.com/methodology/survey/survey-display-499.cfm.

### Definition of variables

#### Outcome variable

The outcome variable was HIV testing among young adult women, a dichotomous variable coded as “1” when a woman reported having “ever been tested for HIV” and “0” when she reported otherwise (never tested for HIV). The information about the dependent variable was generated from this question.

#### Explanatory variables

The explanatory variables used in this study were selected based on the available data in the 2016–2018 PNGDHS datasets and previously published literature. [17,18,41] The independent variables were categorised into socio-demographic factors (age, marital status, education level, occupation, wealth index, place of residence, and region). Age was categorised into “15–19,” 20–24,” and “25–29.” Marital status was recoded into “never married,” “married/living with a partner,” and “previously married.” The occupation was recoded into “not working” and “working.” The wealth indices were determined by household assets owned (e.g., televisions and radio), housing construction materials, and access to water and sanitation facilities [40], and were reported as being “poorer,” “poor,” “middle,” “rich,” and “richer.” A new variable for the wealth index was created and recoded into “poor,” “middle,” and “rich.” Behaviour-related variables comprised the frequency of exposure to mass media (reading a newspaper or magazine, listening to the radio, and watching television), mobile phone ownership, and internet use. Exposure to media was determined by the frequency of reading a newspaper or magazine, listening to the radio, and watching television. Each of these variables had three responses: “not at all,” “less than once a week,” and “at least once a week.” These variables were recoded as “yes” and “no” responses to exposure to media. Use of the internet was identified as “never,” “yes, last 12 months,” “yes, before last 12 months,” and “yes, cannot establish.” The internet use variable was recoded into “no” and “yes.” Furthermore, health risk behaviours include age at first sex, the total number of sexual partners (in the last 12 months), ever being paid for sex (in exchange for cash, gifts, or others), always using a condom during sex, and history of any STI (in the last 12 months). Age at sexual debut was recoded as “less than 20 years” and “20 years or older.” The total number of sexual partners (in the last 12 months) was recoded into “1” and “2 or more.” Incomplete and missing data in the “ever tested for HIV” group remained low (1.2%); therefore, they were excluded from the final analysis.

### Statistical analysis

The final sample data was weighted to restore representativeness and produce a reliable estimate and standard error. Descriptive statistics were used to describe the sociodemographic and HIV-related characteristics of the participants. Bivariate analysis using the Pearson Chi-square (χ^2^) test of independence was performed to examine the relationship between the explanatory variables and the outcome variable. Variables significant at p < 0.25 in univariate analysis were then used in the final multivariable logistic regression model. [42] A Complex Sample Analysis Procedure that accounted for the cluster sampling design and sample weight to provide generalisable and accurate estimates of proportion, probability values, and odds ratios was employed in this analysis. [43,44] The Hosmer-Lemeshow goodness-of-fit was used to test for model fitness. Measures of association were presented as adjusted odds ratios (AORs) with their corresponding 95% confidence intervals (CIs). A *p*-value ≤ 0.05 was used to determine the statistical significance of all analyses. All analyses were conducted using the IBM Statistical Package for the Social Sciences (SPSS), Version 26 (Armonk, NY: IBM Corp.).

### Patient and public involvement

Patients and the public were not involved in the study. We analysed the secondary data available in the public repository.

## Results

### Characteristics of participants

Participants’ demographic characteristics between those who were tested for HIV and those who were not are presented in **Table 1**. Of the total weighted sample of 5,164 young adult women, 3,010 [58.8% (95% CI 57.4%, 60.1%)] were never tested for HIV. The mean age was 21.65 (SD ± 4.23) years. Over half of the women who were not tested for HIV were young (aged < 20 years) (55.1%), never married (79.4%), lacked formal education (63%), not working (60.2%), poor (71.2%), and from rural areas (62.9%). Nearly two-thirds were not exposed to the media (i.e., newspaper or magazine, radio, television). In terms of risky sexual behaviours and related factors, most women who had not tested for HIV reported a sexual debut at age < 20 (61.3%), having one sexual partner (61.7%), having ever been paid for sex (64.9%), always using a condom during sex (51.1%), and not having had an STI in the last 12 months (59.3%). There were statistically significant differences in the proportions of participants who had never been tested for HIV compared to those who had in all the demographic variables, age at sexual debut, the total number of sexual partners, ever being for sex, and having had an STI for the past 12 months in the HIV-related variables (p < 0.05).

**Table 1.** Comparison of participants’ demographic characteristics between those who were ever tested for HIV and those who were not in PNG.

| Characteristics | Participants<br>5,164 (%) | Ever tested for HIV |  | *p*-value* |
| --- | --- | --- | --- | --- |
|  |  | No<br>3,010 (%) | Yes<br>2,154 (%) |  |
| **Age group (years)** |  |  |  | **0.001** |
| 15–19 | 1,880 | 1,035 (55.1) | 845 (44.9) |  |
| 20–24 | 1,725 | 1,028 (59.6) | 697 (40.4) |  |
| 25–29 | 1,559 | 947 (60.7) | 612 (39.3) |  |
| Mean age = 21.65 (SD ± 4.23) | | | | |
| **Marital status** |  |  |  | **< 0.001** |
| Never married | 832 | 661 (79.4) | 171 (20.6) |  |
| Married | 4,005 | 2,182 (54.5) | 1,823 (45.5) |  |
| Divorced/separated | 327 | 167 (51.1) | 160 (48.9) |  |
| **Education<sup>#</sup>** |  |  |  | **0.001** |
| No formal education | 894 | 563 (63.0) | 331 (37.0) |  |
| Primary | 2,652 | 1,535 (57.9) | 1,117 (42.1) |  |
| High/Secondary | 1,422 | 785 (55.2) | 637 (44.8) |  |
| College/Tertiary | 195 | 127 (65.1) | 68 (34.9) |  |
| **Occupation\*** |  |  |  | **< 0.001** |
| Not working | 3,475 | 2,091 (60.2) | 1,384 (39.8) |  |
| Working | 1,620 | 883 (54.5) | 737 (45.5) |  |
| **Wealth Index<sup>#</sup>** |  |  |  | **< 0.001** |
| Poor | 1,977 | 1,407 (71.2) | 570 (28.8) |  |
| Middle | 943 | 540 (57.3) | 403 (42.7) |  |
| Rich | 2,243 | 1,063 (47.4) | 1,180 (52.6) |  |
| **Place of residence** |  |  |  | **< 0.001** |
| Urban | 1,054 | 424 (40.2) | 630 (59.8) |  |
| Rural | 4,110 | 2,586 (62.9) | 1,524 (37.1) |  |
| **Region** |  |  |  | **< 0.001** |
| Southern | 999 | 617 (61.8) | 382 (38.2) |  |
| Highlands | 2,259 | 1,168 (51.7) | 1,091 (48.3) |  |
| Momase | 1,272 | 870 (68.4) | 402 (31.6) |  |
| Islands | 633 | 355 (56.1) | 278 (43.9) |  |
| **Read newspaper/magazine<sup>#</sup>** |  |  |  | **< 0.001** |
| No | 3,311 | 2,160 (65.2) | 1,151 (34.8) |  |
| Yes | 1,834 | 835 (45.5) | 999 (54.5) |  |
| **Listen to the radio<sup>#</sup>** |  |  |  | **< 0.001** |
| No | 3,253 | 2,107 (64.8) | 1,146 (35.2) |  |
| Yes | 1,893 | 896 (47.3) | 997 (52.7) |  |
| **Watch television<sup>#</sup>** |  |  |  | **< 0.001** |
| No | 3,814 | 2,366 (62.0) | 1,448 (38.0) |  |
| Yes | 1,298 | 602 (46.4) | 696 (53.6) |  |
| **Owns a mobile phone<sup>#</sup>** |  |  |  | **<0.001** |
| No | 3,352 | 2,135 (63.7) | 1,217 (36.3) |  |
| Yes | 1,800 | 867 (48.2) | 933 (51.8) |  |
| **Use the internet<sup>#</sup>** |  |  |  | **< 0.001** |
| No | 4,362 | 2,676 (61.3) | 1,686 (38.7) |  |
| Yes | 783 | 325 (41.5) | 458 (58.5) |  |
| **Age of sexual debut (years)** |  |  |  | **< 0.001** |
| < 20 | 3,710 | 2,275 (61.3) | 1,435 (38.7) |  |
| 20 or more | 1,454 | 735 (50.6) | 719 (49.4) |  |
| Mean age = 14.86 (SD ± 8.80) |  |  |  |  |
| **Total number of sexual partners<sup>#</sup>** |  |  |  | **< 0.001** |
| 1 | 3,564 | 2,198 (61.7) | 1,366 (38.3) |  |
| 2 or more | 1,486 | 755 (50.8) | 731 (49.2) |  |
| **Ever being paid for sex<sup>#</sup>** |  |  |  | **0.050** |
| No | 476 | 375 (78.8) | 101 (21.2) |  |
| Yes | 37 | 24 (64.9) | 13 (35.1) |  |
| **Always condom use<sup>#</sup>** |  |  |  | **0.261** |
| No | 1,542 | 761 (49.4) | 781 (50.6) |  |
| Yes | 2,771 | 1,417 (51.1) | 1,354 (48.9) |  |
| **Had any STI (last 12 months)<sup>#</sup>** |  |  |  | **< 0.001** |
| No or don't know | 4,859 | 2,882 (59.3) | 1,977 (40.7) |  |
| Yes | 221 | 85 (38.5) | 136 (61.5) |  |

\* $\chi$ test, $p = 0.05$ . <sup>#</sup>Variables with missing values.

### Factors associated with not testing for HIV

Factors associated with not testing for HIV among young adult women are presented in **Table 2**. In the multivariable logistic regression analysis, those who were never married (AOR: 4.9, 95% CI: 3.6–6.6), had poor wealth index (AOR: 1.8, 95% CI: 1.3–2.5), from rural areas (AOR: 2.0, 95% CI: 1.5–2.6), from the Momase region (AOR: 1.3, 95% CI: 1.0–1.7), did not read newspapers or magazines (AOR: 1.7, 95% CI: 1.3–2.1), did not listen to radio (AOR: 1.5, 95% CI: 1.1–2.0), experienced early sexual debut (< 20 years) (AOR: 1.5, 95% CI: 1.1–1.9), had one sexual partner (AOR: 1.5, 95% CI: 1.2–2.0), and reported no STI in the past 12 months (AOR: 1.8, 95% CI: 1.1–3.1) had higher odds of not testing for HIV. Conversely, young women from the Highlands region had lower odds of not testing for HIV compared with those in the Southern region (AOR: 0.4, 95% CI: 0.4–0.6).

**Table 2.**
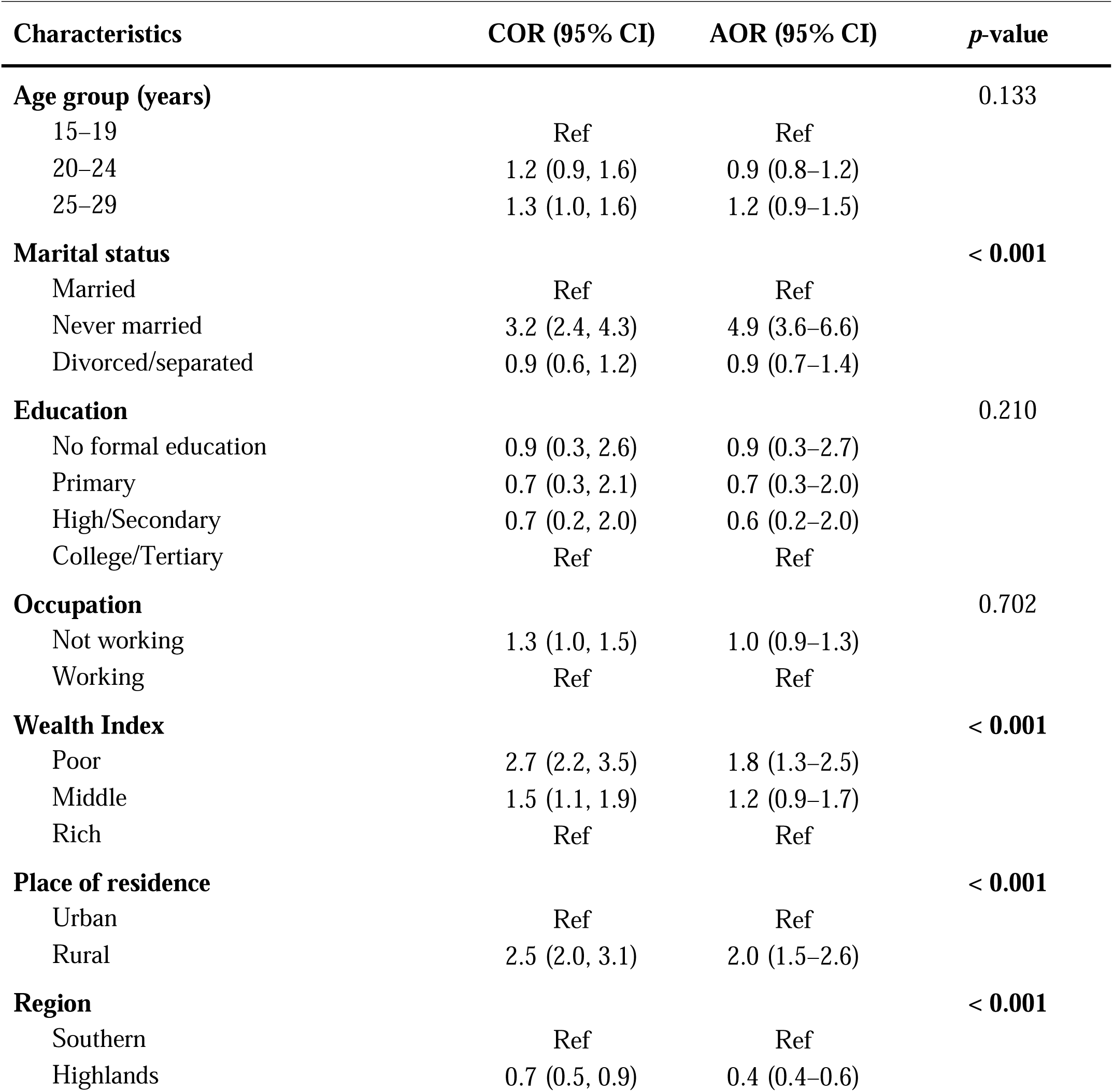
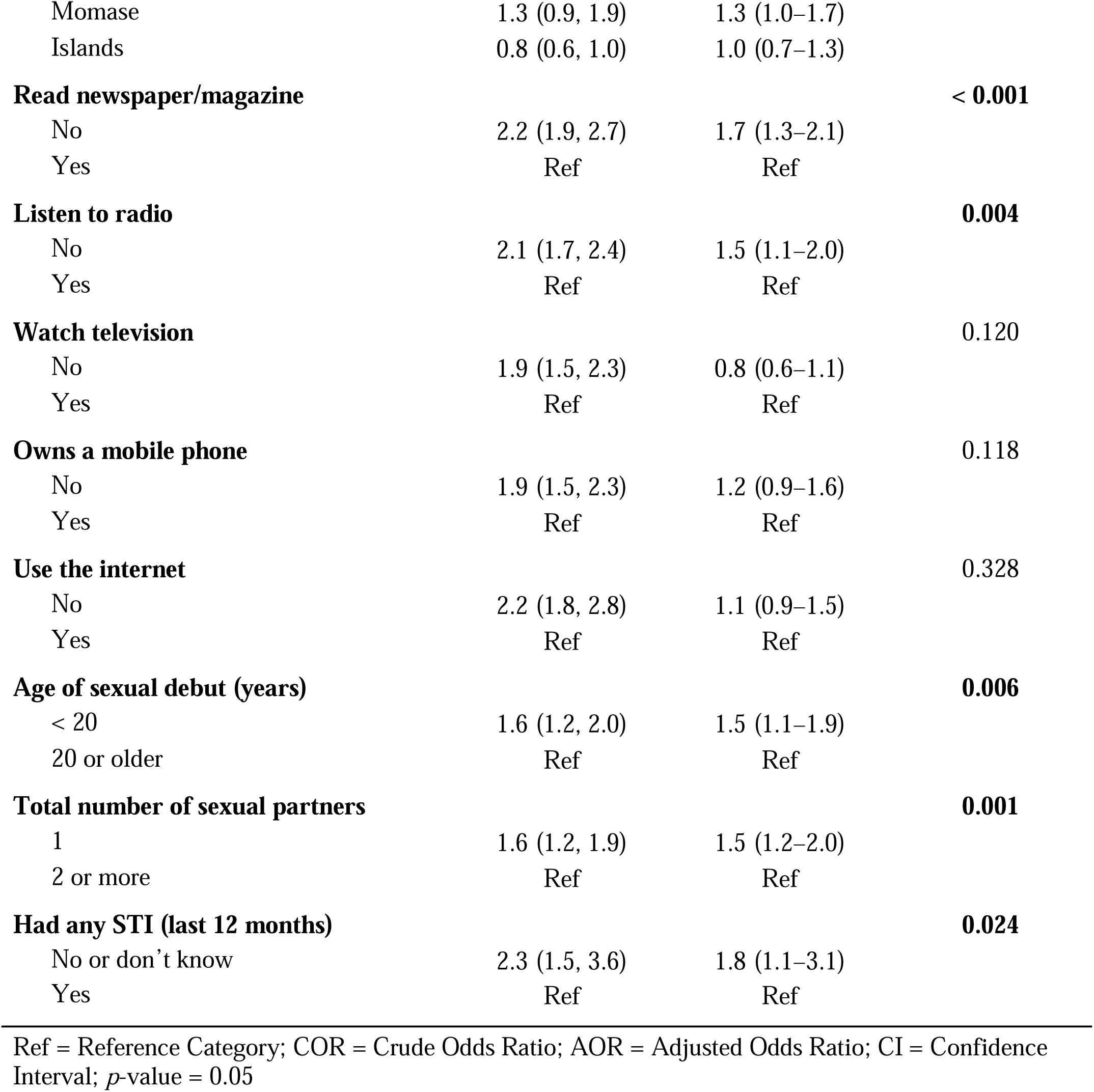
Factors associated with not ever being tested for HIV in the multivariable logistic regression analysis.

| Characteristics | COR (95% CI) | AOR (95% CI) | *p*-value |
| --- | --- | --- | --- |
| **Age group (years)** |  |  | 0.133 |
| 15–19 | Ref | Ref |  |
| 20–24 | 1.2 (0.9, 1.6) | 0.9 (0.8–1.2) |  |
| 25–29 | 1.3 (1.0, 1.6) | 1.2 (0.9–1.5) |  |
| **Marital status** |  |  | < 0.001 |
| Married | Ref | Ref |  |
| Never married | 3.2 (2.4, 4.3) | 4.9 (3.6–6.6) |  |
| Divorced/separated | 0.9 (0.6, 1.2) | 0.9 (0.7–1.4) |  |
| **Education** |  |  | 0.210 |
| No formal education | 0.9 (0.3, 2.6) | 0.9 (0.3–2.7) |  |
| Primary | 0.7 (0.3, 2.1) | 0.7 (0.3–2.0) |  |
| High/Secondary | 0.7 (0.2, 2.0) | 0.6 (0.2–2.0) |  |
| College/Tertiary | Ref | Ref |  |
| **Occupation** |  |  | 0.702 |
| Not working | 1.3 (1.0, 1.5) | 1.0 (0.9–1.3) |  |
| Working | Ref | Ref |  |
| **Wealth Index** |  |  | < 0.001 |
| Poor | 2.7 (2.2, 3.5) | 1.8 (1.3–2.5) |  |
| Middle | 1.5 (1.1, 1.9) | 1.2 (0.9–1.7) |  |
| Rich | Ref | Ref |  |
| **Place of residence** |  |  | < 0.001 |
| Urban | Ref | Ref |  |
| Rural | 2.5 (2.0, 3.1) | 2.0 (1.5–2.6) |  |
| **Region** |  |  | < 0.001 |
| Southern | Ref | Ref |  |
| Highlands | 0.7 (0.5, 0.9) | 0.4 (0.4–0.6) |  |
| Momase | 1.3 (0.9, 1.9) | 1.3 (1.0–1.7) |  |
| Islands | 0.8 (0.6, 1.0) | 1.0 (0.7–1.3) |  |
| **Read newspaper/magazine** |  |  | **< 0.001** |
| No | 2.2 (1.9, 2.7) | 1.7 (1.3–2.1) |  |
| Yes | Ref | Ref |  |
| **Listen to radio** |  |  | **0.004** |
| No | 2.1 (1.7, 2.4) | 1.5 (1.1–2.0) |  |
| Yes | Ref | Ref |  |
| **Watch television** |  |  | 0.120 |
| No | 1.9 (1.5, 2.3) | 0.8 (0.6–1.1) |  |
| Yes | Ref | Ref |  |
| **Owns a mobile phone** |  |  | 0.118 |
| No | 1.9 (1.5, 2.3) | 1.2 (0.9–1.6) |  |
| Yes | Ref | Ref |  |
| **Use the internet** |  |  | 0.328 |
| No | 2.2 (1.8, 2.8) | 1.1 (0.9–1.5) |  |
| Yes | Ref | Ref |  |
| **Age of sexual debut (years)** |  |  | **0.006** |
| < 20 | 1.6 (1.2, 2.0) | 1.5 (1.1–1.9) |  |
| 20 or older | Ref | Ref |  |
| **Total number of sexual partners** |  |  | **0.001** |
| 1 | 1.6 (1.2, 1.9) | 1.5 (1.2–2.0) |  |
| 2 or more | Ref | Ref |  |
| **Had any STI (last 12 months)** |  |  | **0.024** |
| No or don't know | 2.3 (1.5, 3.6) | 1.8 (1.1–3.1) |  |
| Yes | Ref | Ref |  |

Ref = Reference Category; COR = Crude Odds Ratio; AOR = Adjusted Odds Ratio; CI = Confidence Interval; *p*-value = 0.05

## Discussion

This study investigated factors associated with not testing for HIV among young adult women aged 15–29, based on a nationally representative sample. In our analysis of 5,164 young women from the 2016–2018 DHS in PNG, we found that 71% had never tested for HIV. Not testing for HIV was associated with marital status, wealth index, place of residence, region, reading of newspapers and magazines, listening to the radio, age of sexual debut, the total number of sexual partners, and having any STI in the last 12 months preceding the survey, which were significantly associated with not testing for HIV among young adult women. Given the nature of the study, a causal association cannot be established. Also, behavioural desirability bias might have some effect on the underreporting of sexual behaviours since all the responses were self-reported.

Seven out of every 10 young adult women were never tested for HIV, suggesting a significantly low proportion of HIV testing. This finding corresponds with previous studies from sub-Saharan Africa (63.5%) [41], including Ethiopia (66.5%) and Nigeria (76.3%), [17,18] which showed significantly low HIV testing rates among young women. The low HIV testing uptake in the country may be due to differences in HIV testing modalities implemented for young women. Strategies to optimise access to HIV testing need to shift from conventional facility-based testing to community- and home-based testing services. Community and home-based HIV testing have been proven to enhance HIV testing coverage among young women through self-testing and linking them to ART treatment. [45–47] Self-testing for HIV could increase the proportion of individuals who have ever tested, increase testing frequency, and promote early detection and treatment. [48]

In this study, the majority (80%) of women were from poor households and reported to have never been tested for HIV. This finding supports the recent studies showing that young women, particularly in low- and middle-income countries, encounter a heterogeneous risk for HIV based on their household socioeconomic conditions. [49,50] Furthermore, various aspects of socio-cultural structure can potentiate the HIV risk burden for poor women, such as lack of access to information, health care, formal education, economic opportunities, and exposure to violence. [49]

Adolescent girls and young women from economically disadvantaged and poor households are at higher risk for HIV due to their lack of awareness about HIV transmission and their less frequent use of condoms. [49] However, the authors of studies conducted in Malawi and Rwanda found an inverse association between rich households, higher education, and the uptake of HIV testing among young women. [51,52] While earlier studies in PNG have suggested strategies for reducing the gap between socioeconomic quintiles and health among young women, [29] there is still evidence of inequalities. Studies have shown links between HIV testing and better adherence to safe sex practices and women’s empowerment in terms of education, financial independence, control over household financial decisions, and the power of negotiating sex. [53–55] Our findings suggest that there is an urgent need to tailor prevention strategies for HIV for this group that are sensitive to their culture and socioeconomic context. The focus on improving access to HIV testing services should first be on conceptualising gender in a way that is inclusive. Furthermore, there is a need to address barriers related to poor care-seeking behaviours through the provision of youth-friendly services.

Additionally, the findings of this study showed that rural women are twice as likely not to be tested for HIV compared to their urban counterparts, which corresponds with recent studies conducted in Ethiopia [18] and eastern Africa. [56] Low uptake of HIV testing could be due to the difficulty in accessing sexual and reproductive health and HIV services in primary healthcare facilities, including gender issues, societal norms, and a lack of HIV knowledge, as reported in other studies. [27,57,58] Contrary to the findings of our study, the authors of the Ethiopian study have reported an increased uptake of HIV testing among women in rural areas who have higher socio-economic status, non-farming occupations, female-headed households, live within proximity of health facilities, and participate in community dialogues. [59] The utilisation of HIV testing services in rural areas is largely influenced by socio-economic, behavioural, and health service factors.

Our study further found a negative association between the region of residence and not testing for HIV, with women in the Momase region being more likely to not be tested for HIV compared to those in the Southern region. The regional disparities in not testing for HIV in PNG could be a result of the unequal allocation and distribution of health resources, including access to healthcare services [26,31]. This finding highlights the need for interventions aimed at improving HIV testing, especially targeting women from disadvantaged regions of the country.

Women who had no access to at least one media platform (newspapers or radio) were less likely to have been tested for HIV. This confirms the findings from recent Ethiopian studies that a lack of media access and comprehensive HIV knowledge is associated with lower odds of HIV testing. [60,61] It is most likely that young women do not read newspapers or listen to the radio. However, a recent Rwandan study revealed that young women who have access to at least one media platform at least once a week (newspapers, TV, or radio) are more likely to have been tested for HIV. [52] Media exposure increases awareness about the importance of HIV testing and improves understanding of the prevention approach, which might encourage women to be tested for HIV. [60] The results of the study underscore the need to utilise various media platforms in disseminating HIV information to sensitise young women to constantly learn about the benefits of HIV prevention.

Findings from this study showed that the majority (61.3%) of women who debuted sex were aged < 20 and were not tested for HIV compared to their counterparts. The study supports previous findings suggesting a low HIV testing prevalence among this age group in high-prevalence settings. [17] Early sexual debut with risky behaviours has been posited as an important risk factor for HIV infection. Studies have shown that factors such as peer pressure, lower education, coercion, substance use (tobacco and alcohol use), poor parental or guardian attachment, and exposure to pornographic materials were associated with an early sexual debut. [62–64] Furthermore, a lack of HIV knowledge and low-risk perception have been considered a potential barrier to HIV testing among adolescent women. [65] This could be a likely reason why young women were not tested for HIV in this study. The precocious sexual debut predisposes young women to adverse psychological and health outcomes. Thus, the need for timely, age-appropriate, continuous, and enhanced awareness and reproductive health services and interventions is imperative for this priority group.

Regarding the number of sexual partners, women who had one sexual partner had lower odds of being tested for HIV compared with their counterparts. This finding is in contrast with other studies [56], where women with multiple sex partners had a lower chance of being tested for HIV. The low uptake of HIV testing among young women in this study could be due to the perception that they have a lower risk of contracting HIV because they have one sexual partner. Another reason could be that women refuse or forgo testing out of fear of the consequences they may encounter, such as HIV-related stigma and discrimination, including violence from their partners and family for disclosing their HIV-positive status, as reported in previous studies. [27,66,67] There are common notions that female biological vulnerability makes women more likely to contract HIV than men and that women’s socio-economic marginalisation increases their HIV risk. [57] HIV-related stigma can manifest in a variety of ways, including isolation, mockery, condemnation, physical and verbal abuse, and denial of healthcare access, including HIV services. [68] Interventions to address stigma should not be confined to health facilities; instead, they need to be tailored and focus on creating changes in individual knowledge, attitudes, and behaviours rather than broader social and environmental changes.

In this study, participants who had no STIs in the last 12 months were likely not to have ever been tested for HIV, indicating a lack of awareness and knowledge about STIs. Studies have shown that understanding of STIs in terms of acquisition, protection, prevention, and clinical signs and symptoms is limited among young people. [69] Poor knowledge about STI screening and services could be one possible explanation for young women not knowing their STI statuses. Lack of STI information and screening services, acceptability of services, fear of positive test results, fear of invasive procedures, self-consciousness of genital examination, anticipated stigma, and confidentiality concerns have been found to impede STI screening uptake among young people. [70–72] Also, young women may not have access to the required STI information and services to avoid infections or may be hesitant to access the facilities where information is available. This could be another reason for being unsure or not having any STIs. This study highlights the unmet need for STI screening among young adult women in PNG. Urgent efforts are needed to improve STI knowledge through tailored sexual and reproductive health services while taking into account social and cultural norms surrounding gender and sexuality.

### Implications for practice and research

Increasing uptake of HIV testing and counselling among young women can lead to earlier diagnosis, linkage to ART, and retention in care. Interventions that are affordable, feasible, and acceptable for this population to increase access to and the likelihood of being tested remain imperative. To optimise testing coverage, it is necessary to determine young women’s preferences about HIV testing modalities and where they choose to access the service. HIV testing options for young women that include self-testing are promising strategies that may overcome an array of access barriers in this context. In addition, sexual and reproductive health for young women remains crucial to increasing the uptake of HIV testing. There is a need for more sensitisation programmes for healthcare providers, for instance, education and training to sensitise them to reduce HIV-related fear and stigma among young women through user-friendly services. Given that the majority of young women do not seek HIV testing services in PNG, there is a critical need for implementation research on strategies for increasing access to testing, prevention, and treatment. While the findings of this study have provided information about the proportion of young women who have never tested for HIV, it is also important to have an in-depth understanding of the barriers to HIV testing in this age group. Further qualitative research is required to explore barriers and facilitators to HIV testing among this population, which could inform the development of novel HIV prevention strategies.

## Conclusions

HIV testing among young adult women remained low in PNG, with more than half having not been tested for HIV. Factors associated with not testing for HIV included never being married, poor households, rural areas, not having access to media (newspaper, magazine, or radio), early sexual debut, having one sex partner, and an unknown STI status. These factors could allow the epidemic to intensify as young adult women with undiagnosed infections are not likely to be tested for HIV, adopt behaviour change and risk reduction strategies, or access treatment. The use of various media platforms is crucial for disseminating information on HIV/STI prevention, risk reduction, and increasing HIV testing uptake among young women. Efforts to optimise access to HIV testing should also shift from conventional facility-based testing to community- and home-based testing services to target those who are disadvantaged and from rural areas. Furthermore, programmatic approaches from the government, civil society, and communities to overcome barriers associated with the uptake of HIV testing are thus warranted.

## Data Availability

All data produced are available online at the DHS Program

https://dhsprogram.com/data/available-datasets.cfm.

## Acknowledgements

We are grateful to the ICF International and DHS Program for permitting us to use the 2016–2018 PNGDHS dataset to conduct this study.

## Supplementary materials

STROBE Checklist

## Footnotes

### Contributors

MKM conceptualized and designed the study. MKM and FWK conducted statistical analyses. MKM drafted the original manuscript. FWK contributed to the writing and editing of the manuscript. MKM and FWK reviewed and edited the manuscript. All the authors reviewed and approved the final version of the manuscript. MKM is the guarantor and accepts full responsibility for the work.

### Funding

The authors have not declared a specific grant for this research from any funding agency in the public, commercial, or not-for-profit sectors.

### Competing interests

None declared.

### Patient consent for publication

Not required.

### Ethics approval

The 2016–2018 PNGDHS protocol was reviewed and approved by the Institutional Review Board of ICF and Medical Research Advisory Committee of PNG. Informed consent was obtained from all participants before the interviews were conducted. For this study, a formal request to use the raw data was obtained from the DHS program. Permission was granted by the DHS program to access the data, which was anonymized for analysis.

### Provenance and peer review

Not commissioned; externally peer reviewed.

### Data availability statement

The 2016–2018 PNGDHS data are publicly available in a public, open-access repository. Data can be obtained upon a registration-access request from the DHS Program website at https://dhsprogram.com/data/available-datasets.cfm.

## Notes

### Competing Interest Statement

The authors have declared no competing interest.

### Author Declarations

Data is available at DHS Program website: https://dhsprogram.com/data/available-datasets.cfm.

### Summary of Updates

The manuscript has been revised according to the suggestions and comments of the reviewers.

